# Association of Long-Term Blood Pressure Variability with Cerebral Amyloid Angiopathy-related Brain Injury and Cognitive Decline

**DOI:** 10.1101/2024.02.24.24303071

**Authors:** Lukas Sveikata, Maria Clara Zanon Zotin, Dorothee Schoemaker, Yuan Ma, Valentina Perosa, Anthipa Chokesuwattanaskul, Andreas Charidimou, Marco Duering, Edip M. Gurol, Frédéric Assal, Steven M. Greenberg, Anand Viswanathan

## Abstract

**Introduction:** Long-term systolic blood pressure variability (BPV) has been proposed as a novel risk factor for dementia, but the underlying mechanisms are largely unknown. We aimed to investigate the association between long-term blood pressure variability (BPV), brain injury, and cognitive decline in patients with mild cognitive symptoms and cerebral amyloid angiopathy (CAA), a well-characterized small-vessel disease that causes cognitive decline in older adults.

**Methods:** Using a prospective memory clinic cohort, we enrolled 102 participants, of whom 52 with probable CAA. All underwent a 3-tesla research MRI at baseline and annual neuropsychological evaluation over 2 years, for which standardized z-scores for four cognitive domains were calculated. BPV was assessed using a coefficient of variation derived from serial outpatient BP measurements (median 12) over five years. We measured the peak width of skeletonized mean diffusivity (PSMD) as a marker of white matter integrity, and other neuroimaging markers of CAA, including lacunes and cortical cerebral microinfarcts. Using regression models, we evaluated the association of BPV with microstructural brain injury and whether CAA modified this association. We also examined the association of BPV with subsequent cognitive decline.

**Results:** Systolic BPV was dose-dependently associated with PSMD (estimate=0.22, 95% CI: 0.06, 0.39, p=0.010), independent of age, sex, mean BP, common vascular risk factors, brain atrophy, and CAA severity. The presence of probable CAA strengthened the association between BPV and PSMD (estimate=9.33, 95% CI: 1.32, 17.34, p for interaction = 0.023). Higher BPV correlated with greater ischemic injury (lobar lacunes and cortical cerebral microinfarcts) and a decline in global cognition and processing speed (estimate=-0.30, 95% CI: -0.55, -0.04, p=0.022).

**Discussion:** Long-term BPV has a dose-dependent association with alterations in white matter integrity, lobar lacunes, and cortical cerebral microinfarcts, and predicts cognitive decline. Controlling BPV is a potential strategic approach to prevent cognitive decline, especially in early-stage CAA.

**“TAKE-HOME POINTS” FOR SOCIAL MEDIA:**

1. **Twitter handle: @LSveikata**
2. **What is the current knowledge on the topic?** Long-term blood pressure variability (BPV) has been proposed as a novel risk factor for dementia, but the underlying mechanisms are largely unknown. Brains affected by cerebral amyloid angiopathy (CAA), a well-characterized small-vessel disease, may be at risk of developing BPV-related brain injury.
3. **What question did this study address?** Is long-term blood pressure variability (BPV) associated with brain injury and cognitive decline in patients with cerebral amyloid angiopathy (CAA)?
4. **What does this study add to our knowledge?** This prospective memory clinic cohort study demonstrated a dose-dependent relationship between systolic BPV and altered white matter integrity, independent of demographic and vascular risk factors and more pronounced in individuals with evidence of CAA. Higher BPV was also associated with greater ischemic brain injury and cognitive decline.
5. **How might this potentially impact on the practice of neurology?** These findings suggest that BPV may be a modifiable risk factor for brain injury and cognitive decline, particularly in individuals with CAA, and could be targeted in preventative strategies.

## INTRODUCTION

Small vessel diseases (SVD) account for up to 30% of strokes and contribute to up to 50% of dementia cases, but no treatment or effective preventive strategies exist against SVD or cerebral amyloid angiopathy (CAA) specifically, a common and well-characterized SVD and one of the main drivers of cognitive decline in older adults.^1,2^ CAA is defined by amyloid-beta deposits in leptomeningeal and cortical small vessels that cause extensive vessel wall remodeling^3,4^. The severe changes in small vessel wall physiology, including loss of smooth muscle cells, cracking, and fibrinoid necrosis may underlie the impaired vascular reactivity in CAA.^5,6^ Overall, the accumulation of widespread CAA-related ischemic and hemorrhagic injury, together with Alzheimer’s disease, are the most important substrates to dementia in older adults.^1,7^

A diagnosis of probable CAA can be made during life using the Boston criteria with high sensitivity and specificity in patients who present clinical symptoms of CAA, including cognitive impairment.^8,9^ Compelling evidence suggests that intensive blood pressure (BP) control may slow cognitive decline and white matter disease.^10,11^ However, hypertension is not always identified as an independent risk factor, indicating that there may be other underlying feature in BP profiles that influences cerebrovascular outcomes in addition to the mean BP profile.^12^ Visit-to-visit BP variability (BPV), as a marker of long-term BP fluctuations, has emerged as a risk factor for stroke, subclinical white matter disease, cognitive impairment and dementia, with prognostic value beyond that offered by mean BP levels.^12–17^ A post-hoc analysis of SPRINT-MIND trial has reported that participants with the highest BPV were more likely to progress from mild cognitive impairment (MCI) to dementia despite excellent BP control.^16^ The mechanisms underlying this relationship between increased BPV and dementia and whether vascular brain pathology influences this association remain largely unknown.

Early white matter microstructure alterations associated with CAA were shown to be associated with cognitive impairment.^18–20^ Since white matter hyperintensities (WMH) of presumed vascular origin are a strong predictor of dementia, it is crucial to identify and prevent the progression of WMH at an early stage.^21,22^ More advanced imaging metrics, such as the diffusion MRI-based peak width of skeletonized mean diffusivity (PSMD), can detect these alterations even before they are visible on standard clinical imaging.^20,23^ Other markers of CAA, such as cortical cerebral microinfarcts and lacunes, are also strongly associated with cognitive decline.^24–26^ However, the underlying mechanisms behind CAA-related vascular brain injury are not fully understood.

Our hypothesis was that increased BPV may be associated with vascular brain injury due to the underlying CAA-related vessel wall remodeling and alterations in physiological vessel responses to sudden dips and peaks of BP. Using a prospective memory clinic cohort, our objective was to determine if long-term BPV was linked to white matter integrity, brain injury, and domain-specific cognitive decline over time. We also examined whether the presence of CAA imaging markers influenced the relationship between BPV and white matter integrity.

## METHODS

### Study Design

In this single-center prospective memory clinic cohort study, we used a hybrid observational study design, including longitudinally acquired BP data, cross-sectional neuroimaging analysis, and a longitudinal assessment of the association between BPV and cognitive decline, following STROBE reporting guidelines. All research procedures were approved by the Massachusetts General Hospital Institutional Review Board, and written informed consent was obtained from all participants.

### Study Population

Participants with and without CAA presenting mild cognitive symptoms were recruited from the ongoing Massachusetts General Hospital memory clinic cohort between August 2010 and October 2020. Detailed inclusion criteria for cohort enrollment are available in supplemental content, as previously described.^18,27^ In brief, participants with available neuropsychological evaluation, research MRI, and at least 2 BP measures were included in the study. The modified Boston criteria were used to identify probable CAA as described in supplemental content.^28^ These criteria have high specificity and positive predictive value for advanced CAA pathology in hospital-based cohorts presenting cognitive symptoms but no prior intracerebral hemorrhage.^9^ Participants who did not fulfill the probable or possible CAA criteria were classified as non-CAA.

### Data collection and follow-up

Participants underwent baseline clinical evaluation, research MRI, and neuropsychological examination at baseline and annually for two years, as previously described (see supplemental material).^18,27^

### Blood pressure assessments and calculation of BP variability

The BP measurements were collected retrospectively from predetermined study cohort visits and using electronic health records over five years prior to baseline neuroimaging (supplemental content).

In brief, study staff collected information on BP measurements obtained in an outpatient setting by medical personnel over five years prior to baseline imaging, according to previously published methods in cohorts with CAA and cognitive decline.^15,29^ Self-monitoring BP measurements were not considered. The coefficient of variation (CoV) defined as the ratio of the standard deviation to the mean (SD/mean) was used to calculate systolic and diastolic BP variability over five years for every participant. This method is widely accepted for assessing overall BP variability over time and provides more reliable information than SD about fluctuations in BP by correcting for the proportionality observed between the sample mean and variability.^12^

### Neuropsychological Assessment

Global cognition was assessed using the Mini-Mental State Examination ^30^ and composite z-scores adjusted for sex, age, and education were calculated for four cognitive domains using nine individual tests, as previously described in our previous studies (see supplemental content).^18,27^ In brief, we used standardized neuropsychological tests to assess the following cognitive domains: (1) attention/processing speed (Trail Making Test A, Digit Span Forward, Wechsler Adult Intelligence Scale−III Digit Symbol-Coding), (2) executive function (Trail Making Test B and Digit Span Backward), (3) memory (Hopkins Verbal Learning Test– Revised, immediate recall and delayed recall), and (4) language/semantic function (Boston Naming Test and Semantic Fluency, Animals). These summary measures decrease ceiling artifacts and other sources of measurement error.

### Neuroimaging acquisition and analysis

All subjects completed a 3-Tesla research MRI scan (Magnetom Prisma-Fit or TIM-Trio, Siemens Healthineers, Erlangen, German) using a 32-channel head coil. The imaging was performed within 3 months of neuropsychological evaluation (median = 0 days, IQR range 0-62.5). The imaging protocol is detailed in the supplemental content.

Conventional MRI markers of cerebral SVD were rated by an experienced neuroradiologist blinded to clinical information (M.C.Z.Z.), following the Standards for Reporting Vascular Changes on Neuroimaging 2 (STRIVE-2) recommendations, as described previously.^18,26^ In brief, we determined the presence, number, and topography of cerebral microbleeds (CMB), cortical superficial siderosis, the extent of visually-rated white matter hyperintensity (WMH) using Fazekas score, presence and number of lacunar infarcts by region (deep and lobar), and cortical cerebral microinfarcts. Based on imaging markers, a total composite CAA score, that has been previously shown to be associated with CAA-related microangiopathy severity and clinical outcomes, was computed.^31^

To exclude excessive motion artifacts we conducted a thorough visual inspection of diffusor tensor imaging (DTI) as well as a quantitative evaluation using the Total Motion Index proposed by Yendiki et al.^32^ We ran the fully automated PSMD script version 1.5/2020 (http://www.psmd-marker.com), as previously described (Figure 1).^18,20^

**Figure 1.**
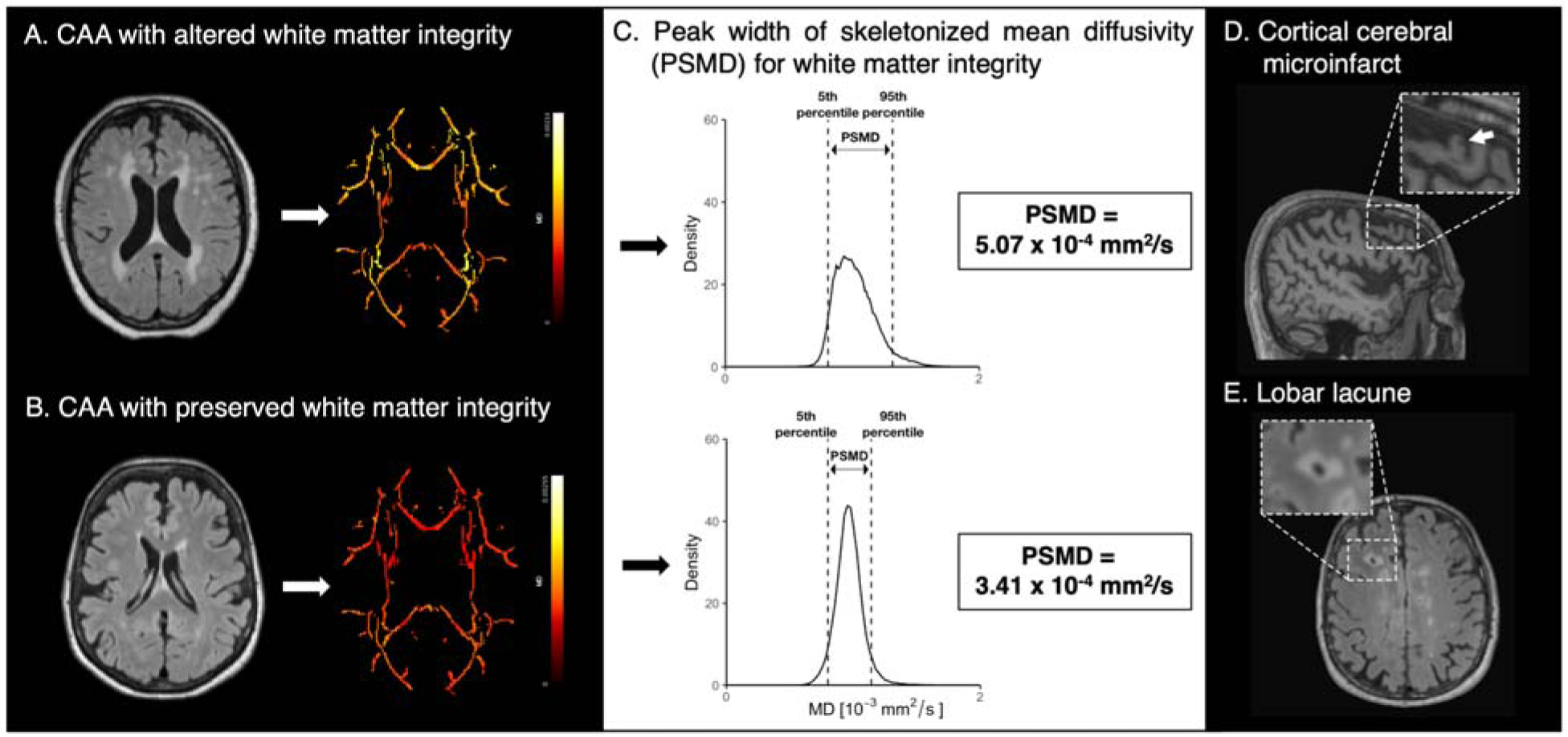
Methods of white matter integrity assessment with peak width of skeletonized mean diffusivity and emerging non hemorrhagic markers in cerebral amyloid angiopathy. (A) Probable cerebral amyloid angiopathy (CAA) subject with high burden of white matter hyperintensity (Fazekas 3) and increased mean diffusivity of main white matter tracts. (B) CAA subject with low white matter hyperintensity burden (Fazekas 1) and preserved mean diffusivity of white main white matter tracts. (C) Histogram analysis allows calculating the peak width of skeletonized mean diffusivity (PSMD). As the PSMD value increases, the integrity of the white matter decreases. Emerging non-hemorrhagic imaging markers of cerebral amyloid angiopathy are (D) cortical cerebral microinfarcts identified on a sagittal T1-weighted images, and (E) lobar lacunes, identified on axial FLAIR images.

The advanced quantitative neuroimaging analysis protocol is described in detail in supplemental material. In brief, we used FreeSurfer (version 6.0) volumetric pipeline to compute the total brain volume, total intracranial volume, and parenchymal brain fraction following a rigorous visual quality inspection, as previously described.^18,27,33^ Following visual inspection for segmentation quality, normalized WMH to intracranial volume (nWMH) was computed using the lesion prediction algorithm^34^ implemented in the toolbox Lesion Segmentation Tool version 3.0.0 for Statistical Parametric Mapping 12.

### Statistical analysis

Descriptive data of demographic, clinical, and radiologic characteristics of participants were reported for the total group and for individuals with and without probable CAA. Logarithmic transformations were used for nWMH volumes and CMB count to achieve a normal distribution. In the primary analysis, we used linear regression models to evaluate the association between BP profile (mean and variability) and PSMD. Multivariable analyses were adjusted for pertinent demographic (age, sex), vascular risk (diabetes, mean systolic BP), and imaging variables (total CAA score and brain parenchymal fraction). These factors were shown to be associated with white matter integrity and cognition and were limited to six covariates to avoid overfitting.^3,23,35^ We reported standardized estimates or odds ratios (OR) with 95% confidence intervals as well as adjusted R^2^ to inform the model fit, where appropriate. An interaction term was used to verify whether the presence/absence of probable CAA modified the association between BPV and white matter integrity. Secondary analyses examined whether there was a dose-effect relationship between BPV and common SVD imaging markers.

We used five models to assess the association between BPV and longitudinal change in cognitive domain-specific performance adjusted for age, sex, and education.

We performed secondary sensitivity analyses, which included adding to the study sample the cases with possible CAA imaging patterns and additionally adjusting the regression models for presence or absence of hypertension, and use of antihypertensive medications. To assess the potential impact of measurement error on our main results, we additionally performed analyses restricting to participants with at least 5 BP measurements.

The proportion of missing data was small (ranging from 0%-2.0% of all the covariates analyzed), and missing data were handled using the missing indicator approach by adding an additional category indicating missing values. Standard diagnostic methods and graphical examination of residuals were used to verify the applicability conditions in regression models. A threshold of p<0.05 was applied for significance. All analyses were performed on R (version 4.1.1).

## RESULTS

We assessed a total of 201 participants from the prospective memory clinic cohort for eligibility and selected 102 eligible non-demented participants (aged 73.4 ± 7.4, 18.6% female) with available data on BP measurements, complete neuroimaging, and cognitive assessments (Figure 2).

**Figure 2.**
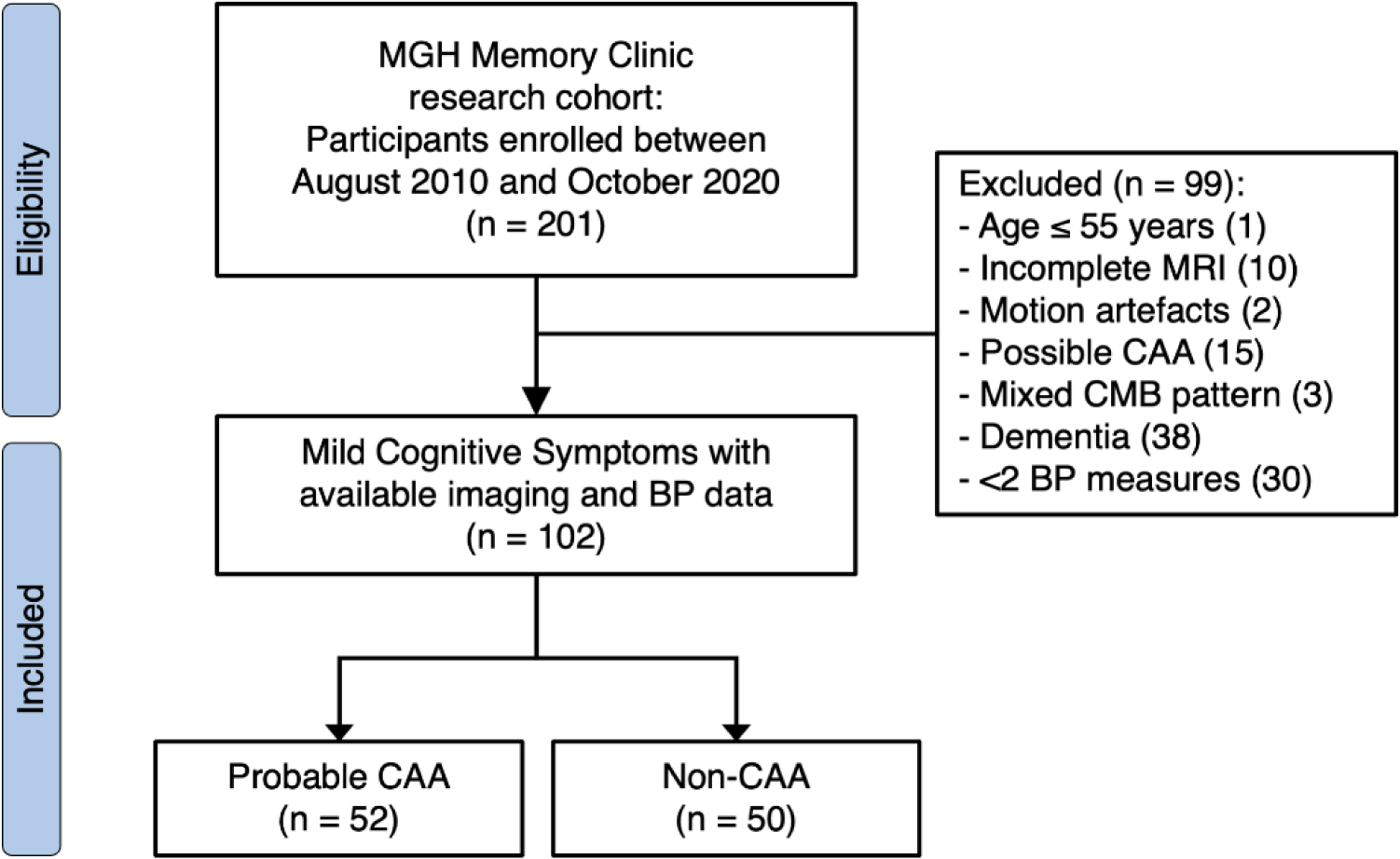
Participant selection flowchart. Abbreviations: BP, blood pressure; CAA, cerebral amyloid angiopathy; CMB, cerebral microbleeds; MGH, Massachusetts General Hospital; MRI, magnetic resonance imaging. Possible CAA was defined as a single CMB in lobar regions or a single cortical superficial siderosis. Mixed CMB pattern was defined by the concomitant presence of lobar and deep microbleeds.

The demographic, clinical, and neuroimaging characteristics of the included study participants are presented in Table 1. Of the total, 52 were identified as probable CAA and 50 as non-CAA. The CAA group had numerically more outpatient BP measurements (median 12 vs. 10 in the non-CAA group, p=0.049). Additionally, the CAA participants had a higher occurrence of cortical cerebral microinfarcts, greater brain atrophy, and higher PSMD values, indicating altered white matter integrity.

**Table 1.**
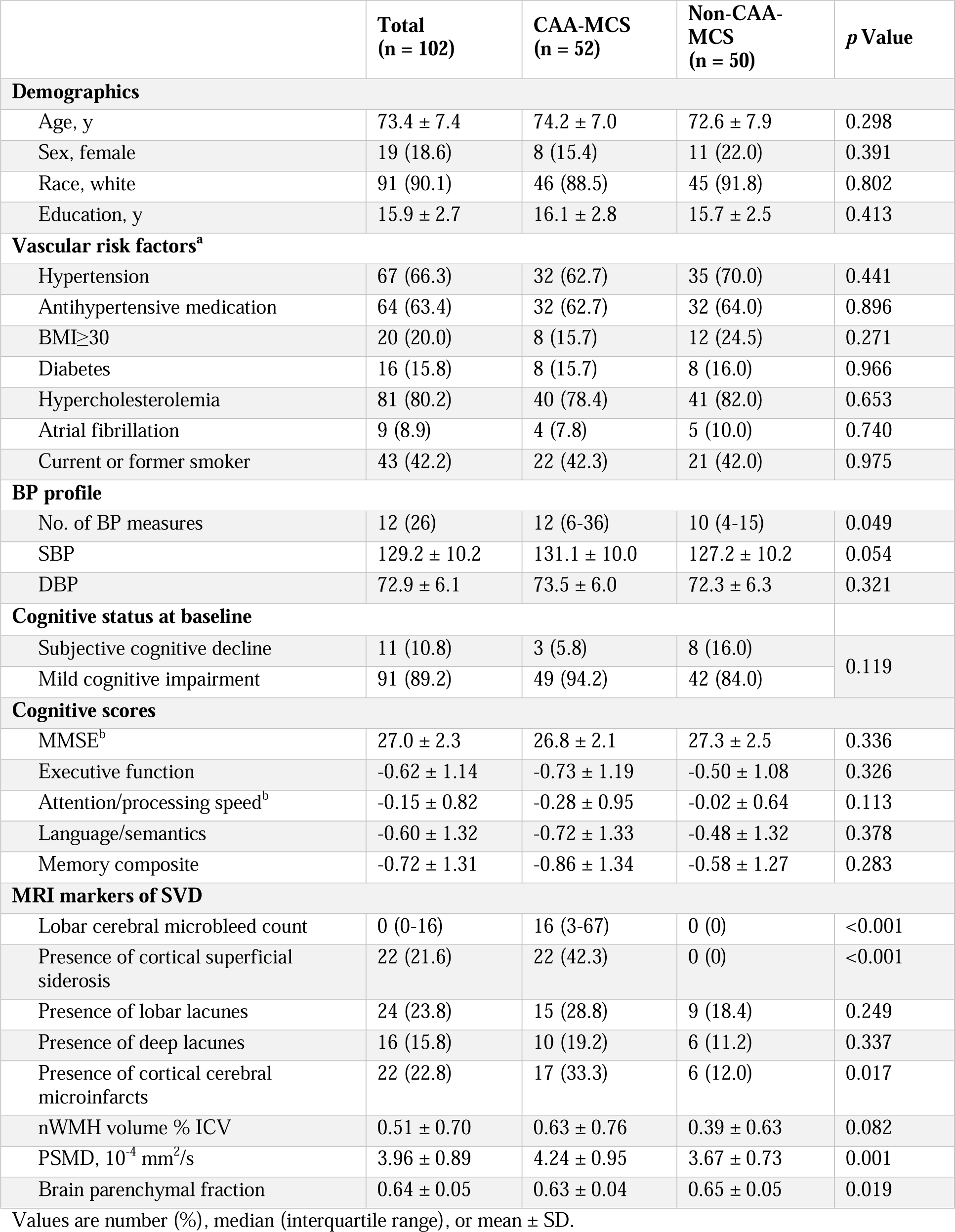
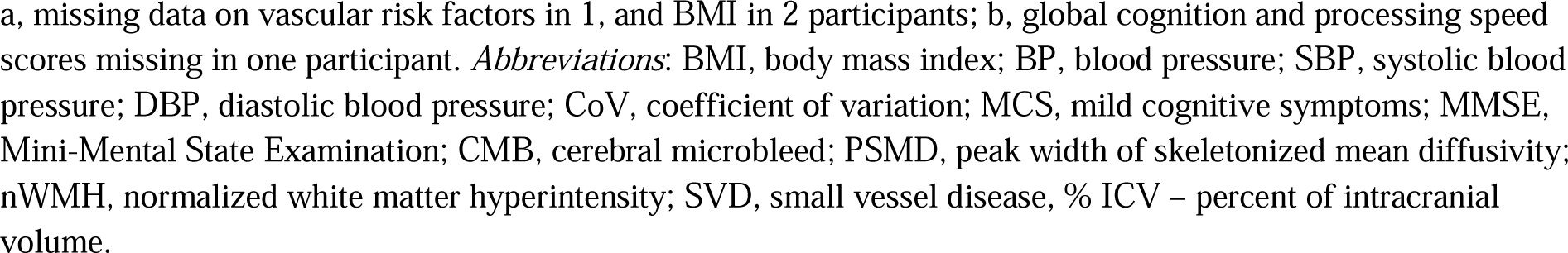
Demographic, clinical, and radiologic characteristics in subjects with mild cognitive symptoms in probable CAA and non-CAA participants.

Hypertension was diagnosed, and antihypertensive medications were administered in 66.3% and 63.4% of participants, respectively. The prevalence of hypertension and treatment regimen for hypertension did not differ between the probable CAA group and the non-CAA group (p=0.667). Among participants taking antihypertensive medication, 29 (45.3%), 25 (39.1%), and 10 (15.6%) used one, two, and three or more drugs, respectively.

### Associations between long-term BP variability and markers of CAA

In linear regression models, we found a strong association between PSMD and systolic BPV (estimate [Est.]=0.39, p<0.001), as well as a moderate association with diastolic BPV (Figure 3B-C). In univariate analysis, PSMD was linked to systolic BPV, mean BP, age, total CAA severity score, and brain volume. However, in a multivariable model, systolic BPV but not mean BP remained a strong predictor of PSMD (Table 2).

**Figure 3.**
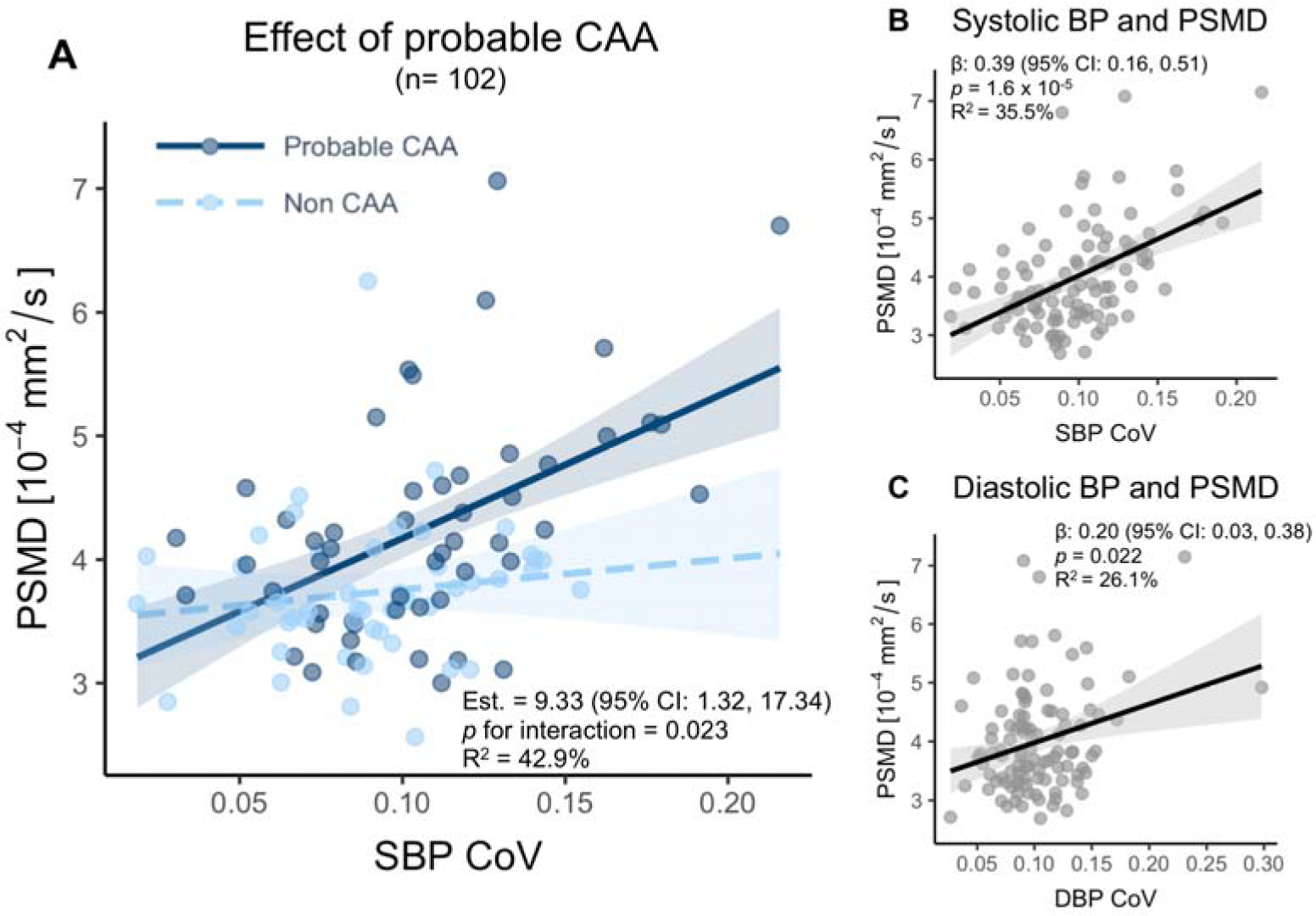
Associations between 5-year blood pressure (BP) variability and white matter microstructural integrity in participants with and without probable CAA. Legend: (A) Interaction plot showing the interaction of probable CAA and non-CAA participants on the association between systolic BP variability and PSMD controlling for age (*PSMD = 0.383 + 0.042*Age + 9.329*[SBP_CV*Probable_CAA])*. Probable CAA increased the effect size of the association between systolic BP variability and PSMD (p for interaction = 0.023). Linear regression models for (B) systolic and (C) diastolic BP variability and PSMD, adjusted for age. Shaded areas represent 95% confidence intervals. Abbreviations: BP, blood pressure; CAA, cerebral amyloid angiopathy; PSMD, peak width of skeletonized mean diffusivity.

**Table 2.** Associations between white matter integrity and blood pressure variability in univariate and multivariable models.

| Variable | Univariate |  | Adjusted<sup>a</sup> |  |
| --- | --- | --- | --- | --- |
|  | Est. (95% CI) | *p* Value | Est. (95% CI) | *p* Value |
| Systolic BP CoV | 0.49 (0.32, 0.67) | <0.001** | 0.22 (0.06, 0.39) | 0.010* |
| Systolic BP mean | 0.25 (0.06, 0.45) | 0.010* | 0.02 (-0.13, 0.18) | 0.790 |
| Age, y | 0.46 (0.29, 0.64) | <0.001** | 0.18 (-0.02, 0.39) | 0.087 |
| Sex, male | -0.01 (-0.21, 0.19) | 0.934 | -0.01 (-0.16, 0.14) | 0.925 |
| Diabetes | 0.14 (-0.04, 0.32) | 0.141 | 0.13 (-0.02, 0.28) | 0.102 |
| Total CAA score | 0.40 (0.22, 0.58) | <0.001** | 0.29 (0.14, 0.44) | <0.001** |
| BPF | -0.49 (-0.66, -0.32) | <0.001** | -0.21 (-0.42, 0.00) | 0.053 |

The primary outcome measure was white matter integrity estimated using peak with of skeletonized mean diffusivity (PSMD). The predictive variables in simple regression models included systolic BP CoV, systolic BP mean, age, sex, diabetes, total CAA severity score, and BPF. All listed covariates were included in the adjusted multivariable model. Abbreviations: BP CoV, blood pressure coefficient of variation; BPF, brain parenchymal fraction; CAA, cerebral amyloid angiopathy; Est., standardized estimate. <sup>a</sup>The variability explained by the multivariable regression model measured by adjusted $R^2 = 39.1\%$ .
\* $p < 0.05$
\*\* $p < 0.01$

### Dose-dependent association between BP variability and CAA imaging markers

We found that systolic BPV was associated with markers of brain injury independent of age, sex, mean BP, and diabetes (Table 3). BPV showed a moderate association with nWMH (Est.=0.22 [0.04-0.41], p=0.021) and a strong association with PSMD (Est.=0.31 [0.11-0.47], p<0.001). We also found that BPV was associated with the presence of cortical cerebral microinfarcts (OR=1.96 [1.10, 3.74], p=0.030) and lobar lacunes (OR=2.03 [1.13, 3.89], p=0.023). The highest quartile compared to the lowest quartile BPV was driving the association with PSMD (Est.=0.72 [0.24, 1.19], p=0.004) and cortical cerebral microinfarcts (OR=7.47 [1.50, 57.27], p=0.024).

**Table 3.** Association between long-term systolic BPV and CAA imaging markers.

| Systolic BPV by quartile |  |  |  |  | Continuous systolic BPV |
| --- | --- | --- | --- | --- | --- |
|  | Q1 | Q2 | Q3 | Q4 |  |
|  |  | *Est. (95% CI)* *p-value* | *Est. (95% CI)* *p-value* | *Est. (95% CI)* *p-value* | *Est. (95% CI)* *p-value* |
| Cerebral microbleed count<sup>a</sup> | Ref. | 0.21(-0.34, 0.77) 0.453 | 0.49 (-0.09, 1.06) 0.101 | 0.22 (-0.37, 0.82) 0.464 | 0.11 (-0.12, 0.33) 0.364 |
| nWMH<sup>a</sup> | Ref. | -0.18 (-0.49, 0.39) 0.442 | -0.04 (-0.25, 0.67) 0.882 | 0.44 (-0.06, 0.93) 0.086 | **0.22 (0.04, 0.41)** **0.021** |
| PSMD | Ref. | -0.05 (-0.52, 0.36) 0.724 | 0.21 (-0.16, 0.77) 0.200 | **0.72 (0.24, 1.19)** **0.004** | **0.31 (0.11, 0.47)** **<0.001** |

|  |  | *OR (95% CI)* *p-value* | *OR (95% CI)* *p-value* | *OR (95% CI)* *p-value* | *OR (95% CI)* *p-value* |
| --- | --- | --- | --- | --- | --- |
| Cortical superficial siderosis | 1.00 | 0.80 (0.19, 3.16) 0.748 | 0.86 (0.18, 3.97) 0.850 | 1.59 (0.38, 6.88) 0.521 | 1.21 (0.69, 2.17) 0.508 |
| Cortical cerebral microinfarcts | 1.00 | 3.78 (0.75, 28.24) 0.132 | 2.97 (0.54, 23.28) 0.234 | **7.47 (1.50, 57.27)** **0.024** | **1.96 (1.10, 3.74)** **0.030** |
| Lobar lacune | 1.00 | 0.37 (0.05, 2.05) 0.277 | 1.23 (0.28, 5.71) 0.786 | 3.09 (0.78, 13.82) 0.119 | **2.03 (1.13, 3.89)** **0.023** |
| Deep lacune | 1.00 | 5.15 (0.60, 113.98) 0.180 | 2.05 (0.22, 45.23) 0.561 | 7.33 (1.06, 148.92) 0.083 | 1.87 (0.97, 3.85) 0.071 |

a, values log-transformed. Data represent standardized estimates (95% CI), odds ratio (OR) values (95% CI) and p-values. Regression models adjusted for age, sex, diabetes, and mean blood pressure (n=102).
*Abbreviations:* BPV, blood pressure variability; CAA, cerebral amyloid angiopathy; PSMD, peak width of skeletonized mean diffusivity; WMH, white matter hyperintensity.
\* p<0.05
\*\* p<0.01

The presence of probable CAA strengthened the association between systolic BPV and PSMD (*p for interaction* = 0.023; Figure 3). Consequently, probable CAA participants, on average, had more severely altered white matter integrity. Our model suggests that a 70-year-old participant with mild cognitive symptoms, systolic BPV in the highest quartile (0.12), and probable CAA would have a PSMD value of 5.60 x 10^-4^ mm^2^/s. This would indicate severely altered white matter integrity, compared to a matched non-CAA subject with a PSMD value of 4.48 x 10^-4^ mm^2^/s, indicating moderately altered white matter integrity.

### Longitudinal cognitive decline and BPV

In patients who had longitudinal neuropsychological testing data (n=59), we found that higher visit-to-visit BPV over five years predicted an annual decline in global cognitive function, especially in processing speed (Figure 4). The association with a decline in processing speed remained significant after adjusting for age, sex, and mean BP (data now shown).

**Figure 4.**
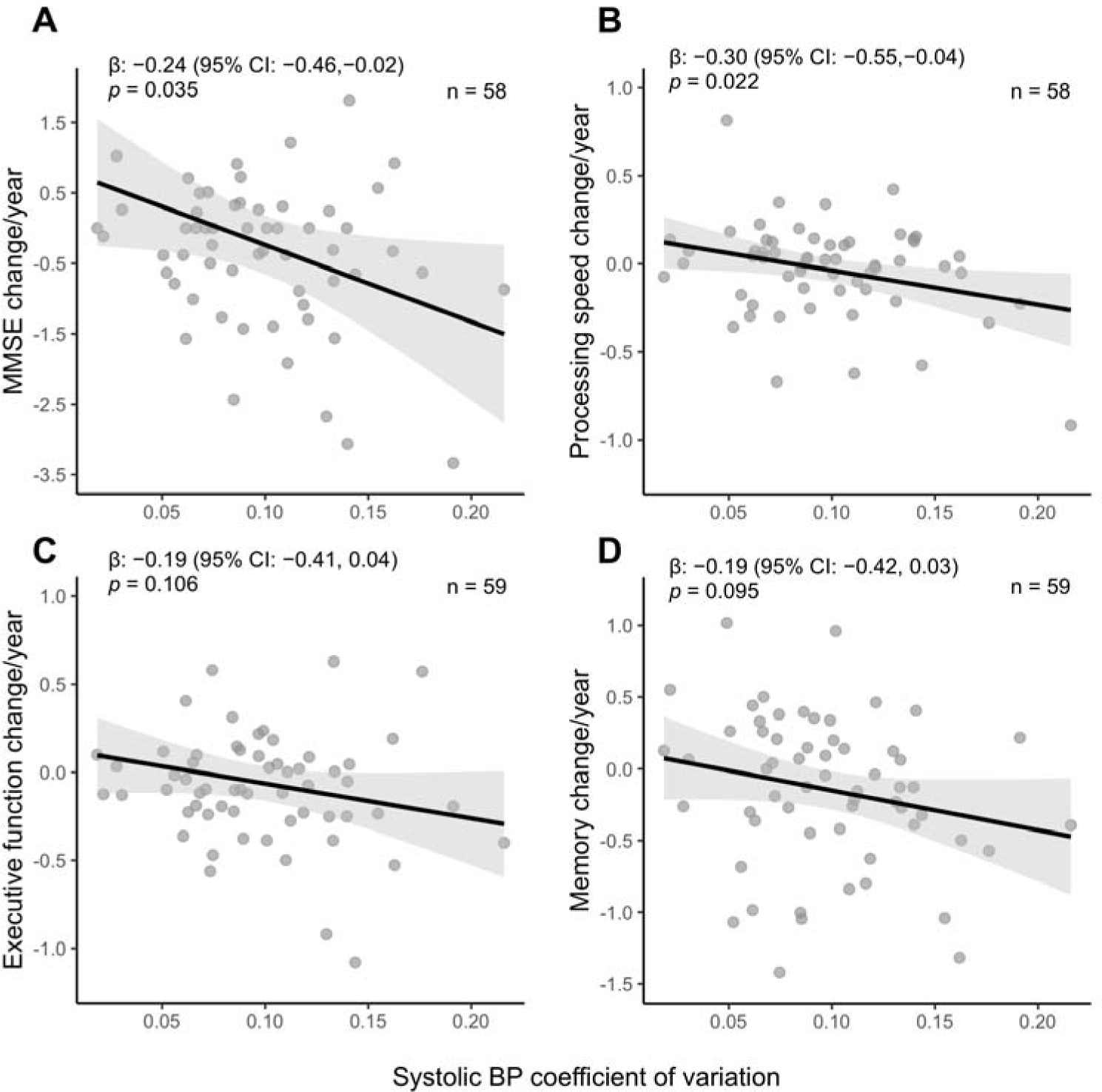
Associations between 5-year BP variability and annualized cognitive decline in non-demented participants with and without probable CAA. Legend: Linear regression models between systolic BP variability and domain-specific cognitive decline in (A) global cognition measured by MMSE, (B) processing speed, (C) executive function, and (D) memory. Cognitive decline is represented in annualized cognitive composite z-scores adjusted for age, sex, and education. Global cognition and processing speed scores were missing in one participant. Associations are presented in standardized β coefficients with shaded areas representing 95% confidence intervals. Abbreviations: BP, blood pressure; CAA, cerebral amyloid angiopathy; MMSE, mini-mental state examination.

### Sensitivity analyses

The excluded non-demented participants were largely similar to the included pool of participants in terms of demographic and clinical characteristics, except for the difference in sex distribution, and slightly higher mean BP (eTable 1). There were no changes in effect sizes in regression models adjusted for sex, history of hypertension, antihypertensive medication use, their number, and frequency of BP measurements (data not shown). The effect sizes remained robust after including participants with possible CAA imaging patterns (n=15). Finally, the association between BPV and white matter integrity was independent of CAA severity and brain atrophy.

## DISCUSSION

In this prospective cohort study of patients exhibiting mild cognitive symptoms, we found that long-term BPV predicted altered white matter integrity, particularly in the presence of probable CAA, independent of age, sex, diabetes, CAA severity, brain atrophy, and mean BP. This suggests BPV as an independent risk factor for vascular brain health. Our findings introduce a new dimension to the understanding of how vascular risk factors interact with CAA-related pathology and contribute to cognitive decline. We also found that increased BPV had a dose-dependent association with lobar lacunes and cortical cerebral microinfarcts, emerging markers of CAA.^25,36–38^ Therefore, our findings suggest that BPV drives silent ischemic brain injury through cerebral microinfarcts, a possible upstream phenomenon related to white matter integrity, and a broader range of CAA-related ischemic brain injury in cortical and subcortical regions.

The debate around whether BPV stems from dysautonomia related to neurodegenerative mechanisms remains unresolved. Although we cannot completely exclude the possibility of reverse causation, the rigorous adjustments for brain atrophy, CAA severity, and longitudinal evaluation of cognitive decline, do not support that BPV was caused by preceding brain injury. Second, our study only included participants with subjective cognitive decline and MCI, and excluded those with mild or severe dementia, thus limiting the possibility that advanced neurodegeneration influenced this association. Further, existing literature does not support the idea that autonomic dysfunction is strongly related to AD and vascular dementia^39^, which makes it less likely that BPV in CAA arises from dysautonomia.

Our data also indicate that elevated BPV could be a risk factor for vascular cognitive impairment, as it predicted domain-specific cognitive decline. We specifically observed a decline in global cognition and processing speed performance, compatible with vascular cognitive impairment phenotype.^3^ In line with our findings, the presence of cerebral cortical microinfarcts has been shown to predict worse cognitive performance.^40^ This might be explained by their perilesional effects on adjacent grey matter and myelin loss in white matter tracts, which affect an area 12 times larger in size than the lesion core.^24^ Our findings add novel insights into the potential drivers of alterations in white matter integrity and brain injury in CAA-affected populations. This study complements previous population-based studies that have generally examined the relationship between BPV and global cognition.^13,15^ We demonstrate that long-term BPV over five years is a predictive factor for clinically detectable cognitive decline. Targeting this potential risk factor for cognitive decline early in the disease course may prevent or slow cognitive decline.

To capture white matter integrity, we employed a quantitative DTI technique known as PSMD.^20,23^ This approach is particularly sensitive to CAA-related changes and has a high repeatability and reproducibility, making it a robust tool for capturing the microstructural brain injuries relevant to vascular brain disease.^18–20,23^ PSMD has been established as a valuable tool for investigating vascular brain injury, as it offers a quantitative measure of the microstructural integrity of principal white matter tracts before changes are visible on conventional imaging (see Zanon Zotin et al.^23^ for a review). Our study adds to a growing body of literature that associates BPV with subclinical brain injury.

We discovered a dose-dependent association between higher BPV and the presence of lobar lacunes and cerebral cortical microinfarcts, considered markers of CAA-related vascular dysfunction.^25,38^ Cortical cerebral microinfarcts and PSMD are emerging as one of the earliest markers of CAA-related brain injury that leads to cognitive decline.^18,23,41^ Interestingly, lobar lacunes and cortical cerebral microinfarcts are not strongly associated with classical vascular risk factors.^24,25,36^ Emerging mechanistic concepts suggest that CAA-related brain lesions stem from vascular dysfunction caused by amyloid deposition decades before clinical manifestations of cerebral hemorrhage and cognitive impairment.^6,37,42^ Pathologic reports of ischemic lesions in advanced CAA have emphasized that the replacement of vascular smooth muscle cells with a rigid ring of beta-amyloid might be responsible for altered vascular physiology.^37,43^ Interestingly, we did not find associations between BPV and hemorrhagic imaging markers of CAA. This supports previous research indicating that microbleeds and microinfarcts are caused by distinct underlying mechanisms, where the latter is related to beta-amyloid-positive cortical vessels.^38^ In line with this, hypoperfusion due to sudden drops in BP has been suggested as an additional potential cause for cortical cerebral microinfarcts^44^, which, as our data suggest, might be further accelerated by impaired small vessel physiology. The current understanding of CAA progression emphasizes that ischemic injury precedes hemorrhagic lesions^37^ and highlights the need for more comprehensive research to elucidate the possible mediatory role of CAA in BPV-induced cerebral microstructural changes.

Additionally, alternative mechanistic explanations may exist, particularly due to improving understanding that both CAA and hypertensive SVD (arteriosclerosis) may interact at the pathophysiological level to cause brain injury.^45^ These two common types of SVD often co-occur, and further studies are needed to disentangle the SVD-type specific effects on brain tissue.

Our findings hold potential implications for preventative strategies aimed at mitigating the risk of dementia. While elevated BP is already an established risk factor, our data suggest that BPV may provide an additional avenue for intervention, particularly in populations susceptible to vascular and mixed dementias. Emerging evidence suggests drug-class-specific effects in reducing BPV, especially in the context of sporadic SVD, though more research is needed to confirm these findings.^17,46^ Given that the CAA-related pathophysiological cascade starts decades before the onset of cognitive decline,^37^ targeting the interventions at the earliest stages of the disease will be essential.

The study results are generalizable to memory clinic populations with mild cognitive symptoms, including subjective cognitive complaints and mild cognitive impairment, although the small group size limits the generalizability to the former population. Our cohort included mostly white individuals with relatively high education levels and only mild cognitive symptoms, as a result limiting the generalizability of the results to populations with different socio-economic statuses, more advanced stages of cognitive decline, e.g., mild dementia, and other clinical settings such as inpatient groups or community-dwelling individuals without cognitive complaints.

The strengths of our study lie in its prospective cohort design, a relatively high number (median=12) of longitudinal BP measurements, and standardized diffusion neuroimaging protocol, enabling us to analyze microstructural brain alterations and domain-specific cognitive associations with relatively smaller sample sizes. Limitations such as the absence of Alzheimer’s disease biomarkers should be acknowledged. This limitation is common in any study related to CAA as it often occurs with Alzheimer’s dementia. Another limitation is the lack of APOE genotype data in our cohort, as according to observational data, e4 allele carriers may be at higher risk of neurovascular dysfunction.^47,48^ CAA participants have a higher prevalence of the APOE e4 allele that maybe one of the underlying factors increasing the association of BPV-related injury in this population. On the other hand, APOE*ε4 is a major risk factor for cognitive decline and vascular amyloid deposition, hence CAA progression.^1^ Therefore, our findings warrant further confirmation in cohorts with APOE genotyping, as these individuals may be most likely to benefit from preventive interventions targeting BPV.

In conclusion, our findings underscore the potential role of BPV in cognitive decline and CAA-related vascular brain injury, reaffirming the urgency to focus on early disease stages for potential therapeutic interventions. Cognitive decline in patients with high BPV may be driven by altered white matter integrity, driven by cortical cerebral microinfarcts and lobar lacunes. Future research should focus on specific antihypertensive treatments that might be most beneficial in controlling BPV, thereby offering a promising avenue for dementia prevention.

## Supporting information

Supplemental Online Content

## Acknowledgments

The authors are thankful to all participants enrolled in the Massachusetts General Hospital memory clinic cohort study. We also thank the Massachusetts General Hospital Stroke Research Center staff for coordination of this study. This work was conducted with support from Harvard Catalyst | The Harvard Clinical and Translational Science Center (National Center for Advancing Translational Sciences, National Institutes of Health Award UL1 TR002541) and financial contributions from Harvard University and its affiliated academic healthcare centers. The content is solely the responsibility of the authors and does not necessarily represent the official views of Harvard Catalyst, Harvard University and its affiliated academic healthcare centers, or the National Institutes of Health.

## Author contributions

L.S., and A.V. contributed to the conception and design of the study. L.S., M.C.Z.Z., D.S., E.M.G. S.M.G., and A.V. contributed to the acquisition and analysis of data. L.S., M.C.Z.Z., D.S., Y.M., V.P., A.C., A.C., M.D., F.A., S.M.G., and A.V. contributed to drafting the text and preparing the figures. All authors made a critical revision of the manuscript for important intellectual content.

## Data availability

Data will be available upon reasonable request to the corresponding author.

