## Supplemental Online Content for "Association of Long-Term Blood Pressure Variability with Cerebral Amyloid Angiopathy-related Brain Injury and Cognitive Decline"

eTable 1. Comparison of clinical, demographic, and radiologic characteristics in included and excluded study participants

|  | Study participants (n = 102) | Excluded non-demented participants (n = 61) | *p* Value |
| --- | --- | --- | --- |
| Demographics | | | |
| Age, y | 73.4 ± 7.4 | 75.5 ± 7.1 | 0.077 |
| Sex, female | 19 (18.6) | 26 (42.6) | <0.001 |
| Race, white | 91 (90.1) | 55 (90.1) | 0.848 |
| Education, y | 15.9 ± 2.7 | 16.4 ± 2.8 | 0.261 |
| Vascular risk factors^a^ | | | |
| Hypertension | 67/101 (66.3) | 45/54 (83.3) | 0.039 |
| Antihypertensive medication | 64/101 (63.4) | 32/54 (59.3) | 0.743 |
| BMI≥30 | 20/100 (20.0) | 13/39 (33.3) | 0.150 |
| Diabetes | 16/101 (15.8) | 13/54 (24.1) | 0.300 |
| Hypercholesterolemia | 81/101 (80.2) | 40/54 (74.0) | 0.500 |
| Atrial fibrillation | 9/101 (8.9) | 3/54 (5.6) | 0.753 |
| Current or former smoker | 43/102 (42.2) | 22/44 (50.0) | 0.488 |
| BP profile | | | |
| No. of BP measures | 12 (26) | 11 (16) | 0.279 |
| SBP | 129.2 ± 10.2 | 134.3 ± 12.6 | 0.024 |
| DBP | 72.9 ± 6.1 | 73.0 ± 5.6 | 0.936 |
| Cognitive status at baseline | | | |
| Subjective cognitive decline | 11/102 (10.8) | 12/61 (19.7) | 0.179 |
| Mild cognitive impairment | 91/102 (89.2) | 49/61 (80.3) |  |
| Cognitive scores | | | |
| MMSE | 27.0 ± 2.3 | 27.5 ± 2.0 | 0.162 |
| Executive function | -0.62 ± 1.14 | -0.32 ± 0.88 | 0.080 |
| Attention/processing speed^b^ | -0.15 ± 0.82 | -0.09 ± 0.49 | 0.605 |
| Language/semantics | -0.60 ± 1.32 | -0.24 ± 1.31 | 0.093 |
| Memory composite | -0.72 ± 1.31 | -0.23 ± 1.30 | 0.022 |
| MRI markers of SVD | | | |
| Lobar cerebral microbleeds | 0 (0-16) | 1 (0-1) | 0.469 |
| cSS presence | 22/102 (21.6) | 4/60 (6.7) | 0.023 |
| Lobar lacune presence | 24/102 (23.8) | 16/60 (26.7) | 0.796 |
| Deep lacune presence | 16/102 (15.8) | 15/60 (25.0) | 0.212 |
| Cortical cerebral microinfarct presence | 22/102 (22.8) | 14/59 (23.7) | 0.758 |
| nWMH volume % ICV | 0.51 ± 0.70 | 0.43 ± 0.56 | 0.485 |
| PSMD, 10^-4^ mm^2^/s | 3.96 ± 0.89 | 4.15 ± 1.15 | 0.259 |
| BPF | 0.64 ± 0.05 | 0.64 ± 0.05 | 1.000 |

Values are number (%), median (interquartile range), or mean ± SD.

a, missing data on vascular risk factors in 1, BMI in 2 participants. *Abbreviations*: BMI, body mass index; BP, blood pressure; BPF, brain parenchymal fraction; SBP, systolic blood pressure; DBP, diastolic blood pressure; MMSE, Mini-Mental State Examination; cSS, cortical superficial siderosis; PSMD, peak width of skeletonized mean diffusivity; nWMH, normalized white matter hyperintensity; SVD, small vessel disease, % ICV – percent of intracranial volume.

Study Population Inclusion/Exclusion Criteria

For the present study, we selected participants who (1) were enrolled in the cohort between August 2010 and October 2020; (2) completed a clinical evaluation, neuropsychological examination, and research MRI; (3) met criteria for subjective cognitive complaints or mild cognitive impairment (MCI) as determined from neurologic and neuropsychological examinations confirming absence or presence of objective cognitive deficits and the absence of functional impairment in activities of daily living;^1^ (6) who had at least 2 BP measures available prior to enrollment; (7) and who fulfilled the modified Boston criteria for probable CAA; they were >55 years old at time of death, had multiple lobar CMBs or one lobar CMB and at least one area of cortical superficial siderosis (cSS).^2^

Exclusion criteria included a history of symptomatic or asymptomatic intracerebral hemorrhage, head trauma, ischemic stroke, CNS tumor, or vascular malformation, vasculitis, and blood dyscrasia; excessive anticoagulation (INR>3.0); evidence of hereditary SVD pathology; incomplete neuropsychological examination and/or research MRI; presence of motion or other artifacts compromising the neuroimaging assessment; primary burden of cerebral microbleeds (CMB) located outside lobar brain regions; unstable medical illness (e.g., unstable angina, advanced renal or liver failure, medication-resistant hypertension). Note that we excluded from the primary analysis participants fulfilling criteria for possible CAA, determined by a single CMB in a lobar region or an isolated presence of cSS. Participants who did not fulfill the probable or possible CAA criteria were classified as non-CAA.

Longitudinal follow-up

Participants underwent baseline clinical evaluation, research MRI, and neuropsychological examination at baseline and annually for two years. The BP measurements were collected retrospectively from predetermined study cohort visits and using electronic health records (EHR) over five years prior to baseline neuroimaging.

Blood pressure assessments and estimates of BP variability

Study staff collected information on BP measurements collected in an outpatient medical setting by medical personnel over five years prior to baseline imaging, according to previously published methods.^3^ Briefly, BP measurements were captured at pre-specified follow-up encounters every 6 months or as part of additional clinical encounters prompted by clinical needs. Additionally, we screened EHR for at-home BP measurements taken by a visiting medical professional. Self-monitoring BP measurements were not considered. As in prior studies, we have excluded measurements obtained in inpatient settings (including emergency departments) to avoid capturing short-term BP variability attributable to acute medical events. For all encounters, BPs were measured using an automated BP machine with the patient in a sitting position after a five-minute rest. For all measurements, two BP readings were obtained, and the average BP was recorded. Only measurements that included the value of both systolic and diastolic BP were considered eligible for analysis.

The coefficient of variation (CoV) was used to calculate systolic and diastolic BP variability for each participant. CoV was defined as the ratio of the standard deviation to the mean (SD/mean) of the subjects’ BP measures over five years. This method is widely accepted for assessing overall BP variability over time and provides more reliable information than standard deviation about fluctuations in BP by correcting for the proportionality observed between the sample mean and variability.^4^

Clinical and Neuropsychological Assessment

Medical history and clinical neurological status were assessed for each participant by a neurologist at baseline and follow-up. The clinical assessment included documentation of vascular risk factors, including hypertension, BP measurement as described above, antihypertensive drug usage, body mass index, diabetes, hypercholesterolemia, atrial fibrillation, and smoking status.

Global cognition was assessed using the Mini-Mental State Examination (MMSE).^5^ Using standardized neuropsychological tests, we assessed the following cognitive domains: (1) attention/processing speed (Trail Making Test A, Digit Span Forward, Wechsler Adult Intelligence Scale−III Digit Symbol-Coding)^6,7^, (2) executive function (Trail Making Test B and Digit Span Backward), (3) memory (Hopkins Verbal Learning Test–Revised, immediate recall and delayed recall)^8^, and (4) language/semantic function (Boston Naming Test and Semantic Fluency, Animals)^9,10^. Performance on each test was transformed into sex-, age-, and education-adjusted z-scores. These summary measures decreased ceiling artifacts and other sources of measurement error and have been used in our previous studies.^11–15^ All data were reviewed by a neuropsychologist, masked to clinical variables and previous cognitive measures.

Diagnosis of subjective cognitive decline, MCI, and dementia were based on neurological and neuropsychological examinations, according to the *‘Diagnostic and Statistical Manual of Mental Disorders, 5th edition’* and defined as the presence of a cognitive complaint without evidence of clear decline on neuropsychological tests for subjective cognitive decline, objective decline in at least one domain without impairment in daily life activities for MCI and as objective and progressive cognitive decline, as the reason for impairment in daily life activities, for dementia.^16,17^

Neuroimaging acquisition

All subjects completed a 3-Tesla research MRI scan (Magnetom Prisma-Fit or TIM-Trio, Siemens Healthineers, Erlangen, German) using a 32-channel head coil at Massachusetts General Hospital. The imaging was performed within 3 months of neuropsychological evaluation (median = 0 days, IQR range 0-62.5). The protocol included a 3-dimensional (3D) T1-weighted (repetition time [TR] = 2300-2510 ms; echo time [TE] = 1.69-2.98 ms; resolution = 1 x 1 x 1mm), a 3D fluid-attenuated inversion recovery (TR = 5000-6000; TE = 356-455 ms, resolution = 1 x 1 x 1mm), a susceptibility-weighted imaging (TR = 27-30 ms, TE = 20 ms, slice thickness = 1.4 mm, in-plane resolution = 0.9 x 0.9mm), and a 3D multi-directional diffusion-weighted imaging (60-64 directions, TR = 8000-8040 ms, TE = 82-84 ms, resolution = 2 x 2 x 2mm, b-value = 700 s/mm^2^).

Conventional SVD neuroimaging markers

Conventional MRI markers of cerebral SVD were rated by a neuroradiologist blinded to clinical information (M.C.Z.Z., 9 years of experience), following the Standards for Reporting Vascular Changes on Neuroimaging 2 (STRIVE-2) recommendations, as described previously.^13,18^ In brief, we determined the presence, number, and topography of CMB, cSS, the extent of visually-rated white matter hyperintensity (WMH) using Fazekas score, presence and number of lacunar infarcts by region (deep and lobar), and cortical cerebral microinfarcts (Figure 2). Based on imaging markers, a total composite CAA score, that has been previously shown to be associated with CAA-related microangiopathy severity and clinical outcomes, was computed.^19^

PSMD processing

To exclude excessive motion artifacts we conducted a thorough visual inspection of DTI images as well as a quantitative evaluation using the Total Motion Index proposed by Yendiki et al.^20^ We ran the fully automated PSMD script version 1.5/2020 (<http://www.psmd-marker.com)>, as previously described (Figure 2).^13,21^

Advanced quantitative neuroimaging analysis

We used FreeSurfer (version 6.0) volumetric pipeline to compute the total brain volume (TBV; i.e., brain segmentation volume), and the estimated total intracranial volume (ICV), following a rigorous visual quality inspection, as previously described.^13,22^ To obtain an estimation of global brain atrophy, the brain parenchymal fraction (BPF) was calculated by computing the ratio of the total brain volume to the total intracranial volume, as described previously. ^11^

Following visual inspection for segmentation quality, WMH volume was computed using the lesion prediction algorithm^23^ implemented in the toolbox Lesion Segmentation Tool version 3.0.0 for Statistical Parametric Mapping 12. The optimal threshold of 0.5 was defined as previously described.^13^ To account for variation in brain sizes, the volume of white matter hyperintensity was normalized (nWMH) to ICV: (white matter hyperintensity volume/ICV) × 100.

Statistical analysis

Descriptive data of demographic, clinical, and radiologic characteristics of participants were reported for the total group and for individuals with and without probable CAA. Logarithmic transformations were used for nWMH volumes and CMB count to achieve a normal distribution. In the primary analysis, we used linear regression models to evaluate the association between BP profile (mean and variability) and PSMD. Multivariable analyses were adjusted for pertinent demographic (age, sex), vascular risk (diabetes, mean systolic BP), and imaging variables (total CAA score and brain parenchymal fraction). These factors were shown to be associated with white matter integrity and cognition and were limited to six covariates to avoid overfitting.^24–26^ We reported standardized estimates or odds ratios (OR) with 95% confidence intervals as well as adjusted R^2^ to inform the model fit, where appropriate. To test whether probable CAA modified the effect of BP variability on white matter integrity, we used an interaction term between systolic BPV and the presence/absence of probable CAA. Secondary analyses examined whether there was a dose-effect relationship between BPV and common SVD imaging markers. We used regression models with systolic BPV quartiles as predictors of CMB count, nWMH volume, PSMD, presence of cSS, cortical cerebral microinfarcts and lobar/deep lacunes as outcomes in separate regression models, adjusted for age, sex, diabetes, and mean systolic BP. The lowest quartile BPV was used as a reference value.

Because cognitive function is a complex construct with a range of functions (domains) and levels of function and impairment, we used five models to evaluate the association between BPV and longitudinal cognitive change. We used linear regression models with global cognition and four separate models with specific cognitive domains as outcomes. The cognitive composite scores were represented as annualized changes in global cognition and cognitive domain composite scores computed from z-scores adjusted for age, sex, and education.

We performed secondary sensitivity analyses, which included adding to the study sample the cases with possible CAA imaging patterns and additionally adjusting the regression models for presence or absence of hypertension, and use of antihypertensive medications. To assess the potential impact of measurement error on our main results, we additionally performed analyses restricting to participants with at least 5 BP measurements.

Standard diagnostic methods and graphical examination of residuals were used to verify the applicability conditions in regression models. A threshold of p<0.05 was applied for significance. All analyses were performed on R (version 4.1.1).
